# Clinical implication of time of stroke among post stroke survivors from eastern India: A circadian perspective

**DOI:** 10.1101/2024.07.10.24310104

**Authors:** Dipanwita Sadhukhan, Arunima Roy, Tapas Kumar Banerjee, Prasad Krishnan, Piyali Sen Maitra, Joydeep Mukherjee, Kartick Chandra Ghosh, Subhra Prakash Hui, Arindam Biswas

**Author notes:** <u>Corresponding author</u>: Dr. Dipanwita Sadhukhan, Molecular Biology & Clinical Neuroscience Division National Neurosciences Centre, Calcutta, Peerless Hospital (2^nd^ floor) 360, Panchasayar, Kolkata 700094 India, Dr. Arindam Biswas, Molecular Biology & Clinical Neuroscience Division National Neurosciences Centre, Calcutta, Peerless Hospital (2^nd^ floor), 360, Panchasayar, Kolkata 700094 India.

## Abstract

**Introduction:** The circadian variation in stroke occurrence is a well-documented phenomenon. However, the circadian effect on stroke outcome, particularly on post-stroke cognition, is not yet been fully elucidated.

**Aims and objective:** We aim to evaluate the influence of diurnal variation of stroke onset upon post-stroke cognition and development of post-stroke depression.

**Materials and Methods:** Based on 4-hourly time period of stroke occurrence, 249 recruited cohorts were categorized into 6 groups. Several clinical and cognitive parameters were compared among the groups. Then, the mRNA expression of core clock genes in Peripheral Blood Mononuclear Cells were quantified and correlated with post stroke outcome among 24 acute phase cases with day-time or night-time stroke occurrence. Furthermore, the genetic susceptibility towards higher number of cases in morning were examined by genotyping *CLOCK* (rs1801260T/C, rs4580704G/C) and *CRY2* (rs2292912C/G) genes variants in cases and 292 controls.

**Results and conclusion:** Among the six subgroups, the subgroup-1 represents the nocturnal-onset stroke cases; they were identified to have higher NIHSS score (12.2±5.67) at the time of admission than other subgroups (8.73±5.92). In the same cohort, more number of diabetic individuals with higher fasting blood sugar level (186.57±93.40) was also observed. After 6 months, those with nocturnal-onset stroke had higher prevalence of language impairment and depression score. A significant decrease in mRNA level of *BMAL1* and *CRY1* genes correlated with raw score for language and depression; data in nocturnal onset cases further established our observation. However, the higher incidence of stroke in day time did not reveal any genetic correlation.

## Introduction

The incidence of stroke demonstrates a circadian variation in human beings. For both the subtypes, major peak of occurrence is in day time while minor peak is observed during night. However, the relation between time of stroke onset with prognosis has not yet been fully elucidated. A study from Japan, concluded morning strokes as more likely to be fatal than afternoon strokes [1]. On the other hand, in a recently published Korean study (n = 17,461), nocturnal-onset strokes were identified as more severe and were associated with poor prognosis compared to daytime-onset strokes independent of other covariates and revascularization therapy [2]. At laboratory level, experimental studies indicated that night time injury (active phase) in mice to be associated with less neuronal death due to alteration in circadian clock genes than day time (inactive phase) [3].

These circadian genes tightly control a number of vascular risk factors (hypertension, platelet aggregation, etc.), cellular (microglial activation, angiogenesis, etc.) and molecular events (neuroprotection, neuroinflammation, etc.) which are related to cause and consequences of stroke. It is evident from animal studies that a higher amplitude of misalignment of circadian rhythm is associated with poor stroke recovery through dysregulation of immune responses. However, only a single study in human subjects is reported describing loss of rhythmicity of circadian genes in five ICH patients [4]. In our previous study, we have shown that the genetic variants in *CLOCK, CRY2* and *BDNF* as risk factors for Post-stroke cognitive impairment (PSCI) among eastern Indians [5]. At the same time, a lowering in expression of *CLOCK* and *BDNF* genes in PSCI patients than controls was also described as underlying mechanism for post-stroke cognitive decline. Likewise, such altered expression of circadian genes also has been reported in patients with depression [6]. However, the circadian attributes in post stroke depression development yet to be evaluated.

Since BMAL1 is a key protein in circadian rhythm and has a protective effect on neuronal apoptosis after stroke and was reported to be dysregulated in depression, here we hypothesise that, “changes in temporal profile of circadian genes’ expression during acute phase of stroke may be time dependent and therefore the time of stroke onset can also be a predictor of long-term cognitive outcome and Post-stroke Depression (PSD).”

Therefore, given the background, here we aim to investigate a) a correlation between time of stroke onset to the cognitive status and depression among post stroke survivors; b) expression profile of circadian genes in peripheral blood cells of stroke patients with different injury time; c) genetic susceptibility conferred by clock genes as a reason for major and minor peak of stroke observed in our study cohort.

## Materials and Methods

### Patients and Controls

The present study was conducted upon 253 stroke patients recruited from the National Neurosciences Centre Calcutta and Nilratan Sircar Medical College & Hospital, Kolkata, India. The Ethics Committee of above-mentioned Institutes approved the study protocol. Informed consent was taken as per guidelines of the Indian Council of Medical Research, National Ethical guidelines for Biomedical and Health research involving human participants, India. All the cohorts were adults with neuroimaging evidence of acute stroke. Of all, ∼42% of cases account of small vessel disease. All had Bengali as their mother tongue, able to comprehend and speak, and able to read and write with minimum education level 10^th^ standard. Exclusion criteria include cancer patients, cases suffering from liver, kidney and endocrine diseases, other neurological disorders such as Alzheimer’s disease, Parkinson’s disease, severe dementia without history of stroke, cases with dysphasia or any other form of language deficit, significant psychiatric illness, substance abuse, and history of psychotropic drug intake, drugs known to cause cognitive impairment and major head injury. The demographic details of patients are summarized in Table 1. The history of known risk factors for stroke including hypertension, diabetes mellitus, hyperlipidaemia, and ischemic heart disease was collected from the patients’ medical files, but data on alcohol consumption or smoking were collected while conversing with patient or respective caregiver. A total of 292 individuals as age– and ethnicity-matched unrelated healthy controls with no personal or family history of stroke or any neurological symptoms were recruited in the present study from Kolkata.

**Table 1:** Demographic details of study subjects.

| **Demographic details / subjects** | **Cases (n = 253)** | **Controls (n = 11)** |
| --- | --- | --- |
| Age range | 24 – 83 | 54 - 76 |
| Age at onset ± SD | 54.81 ± 12.30 | Not applicable |
| Male | 210 (83%) | 10 (91%) |
| Female | 43 (17%) | 1 (9%) |
| Family history of stroke | 121 (47.8%) | 0 |
| Hypertension | 197 (77.9%) | 1 (9%) |
| Diabetic | 106 (41.9%) | 3 (27%) |
| Previous known heart disease | 18 (7.1%) | 0 |
| Smoking/Alcohol | 136 (53.8%) | 4 (36%) |
| High blood cholesterol (> 200 mg/dl) | 24 (9.5%) | 2 (18%) |
| High blood triglyceride (> 150 mg/dl) | 28 (11.07%) | 1 (9%) |
| BMI ± SD | 22.72 ± 2.76 | 22.3 ± 2.71 |
| NIHSS ± SD (at admission) | 8.80 ± 5.92 | Not applicable |
| Recurrent event of stroke | 72 (28.5%) | Not applicable |
| Death after stroke | 32 (12.6%) | Not applicable |

In addition, the time of incidence of stroke was recorded for patients for categorisation into six subgroups-based on onset time: subgroup 1 (0:00 – 03:59); subgroup 2 (04:00 – 7:59); subgroup 3 (08:00 – 11:59); subgroup 4 (12:00 – 15:59); subgroup 5 (16:00 – 19:59) and subgroup 6 (20:00 – 23:59). Here, all enrolled study subjects had no previous record of sleep disturbances or recent travel across different time zones.

### Cognitive Parameters and Depression scoring

Cognitive assessment was carried out within 6 to 12 months of stroke onset. The evaluation was done as mentioned in Sadhukhan et al., 2023 [5]. In brief, comprehensive cognitive tests for attention, language, memory, visuospatial skill, executive functions, etc. were analysed in detail using Kolkata Cognitive Screening Battery for primary assessment [7]. Post-stroke depression (PSD) assessment was done using Geriatric Depression Scale (GDS) within 6-12 months of stroke event.

### Selection of SNVs

On basis of a) association of following SNVs like rs1801260C/T, rs4580704G/C of *CLOCK* and rs2292912C/G of *CRY2* with PSCI among eastern Indians as well b) evidence of functional role of these SNVs from experimental data, eQTL (expression quantitative trait locus) findings reported on the GTEx portal website (www.gtexportal.org) and rSNP base prediction (http://rsnp.psych.ac.cn/quickSearch) and here we genotyped these variants to evaluate their association with the circadian pattern of stroke onset. Additionally, *APOE* genotypes (E2/E3/E4) were also determined in study cohort.

### Genomic DNA Isolation and genotyping of SNVs

Genomic DNA isolation and genotyping of selected variants was done as previously described in Sadhukhan et al., 2023 [5]. Briefly, isolation of DNA from peripheral blood by salting out method was followed by polymerase chain reaction (PCR) using the specific primer pairs and restriction digestion of the amplicon using suitable enzymes (New England Biolabs Inc., Ipswich) following manufacturers’ protocol. About 10% of the samples for each variant were randomly selected for sequencing to rule out genotyping errors.

### Determination of mRNA level

The blood samples were collected from stroke patients within 48 hours of stroke onset. The blood was drawn from both patients and controls between 9 AM to 11 AM to control diurnal variation in circadian gene expression. Isolation of peripheral blood mononuclear cells (PBMCs) using Histopaque 1077 (Sigma-Aldrich, St. Louis, US) double-gradient density centrifugation was followed by total RNA extraction using Trizol reagent (Thermo Fisher Scientifc, US). The transcripts level of CLOCK, BMAL1, PER1 and CRY1 expression were quantified by real-time PCR (qPCR) using cDNAs synthesised from equal amount of RNA (2 µg) from each sample as described in our previous study. The gene specific primers were as follows: *CLOCK*_F: 5′-GGGGCAGTCATGGTACCTAG-3′; *CLOCK*_R: 5′-GCCTGAGATGGTTGCTGAAC-3′; *BMAL1*_F: 5′-TGAAGACAACGAACCAGACA-3′; *BMAL1*_R: 5′-CGTGCCGAGAAACATATTCC-3′; and *CRY1*_F: 5′-GGAAAGGAACGAGACGCAGC –3′; *CRY1*_R: 5′-AGTTAGAGGCGGTTGTCCAC –3′ designed with the web-based software Primer3. For, normalization purpose, expression of 18S used as endogenous control. Here, the Quant Studio 5 Real-Time PCR (Applied Biosystems, US) instrument was programmed for quantitation of C_t_ values for each gene. However, for technical failure here we could not amplify the PER1.

## Statistical Analysis

Hardy-Weinberg equilibrium (HWE) at the polymorphic sites was tested using a chi-square test with one degree of freedom. Genotype association was evaluated for p-value, odds ratio and 95% confidence interval (CI) using Javastat (http://statpages.info/ctab2x2.html). To identify circadian variation for stroke onset in our dataset, we tested a null hypothesis stating “there is a lack of circadian variation for IS stroke onset during a day” by using online Chi square calculator for goodness of fit at a significance level p ≤ 0.05. Here, we first assumed that frequency of stroke onset is equal all throughout the 24 hours of the day and therefore compared the observed number relative to the proportion expected, based on the total number of cases for each stroke type [8].

Statistical analysis for gene expression data was performed in Graph Pad Prism 5.0 using Mann-Whitney U-test to compare cases and controls. Two-tailed test was done with a significance level p < 0.05. Data presented as mean log_2_ transformed expression ± SEM. Correlation analyses between *BMAL1, CRY1, CLOCK*, raw score for language and GDS, were performed by Spearman correlation analysis.

## Results

### Demographic characteristics of study subjects

In the present study, demographic data and the principal variables for Ischemic (IS) cases were represented for 253 cases and 11 age, ethnicity matched control individuals as shown in Table 1. Since the control individuals were only recruited for RNA study, the traditional stroke risk factors (like hypertension, diabetes, previous history of heart disease, habit of smoking/alcohol, high blood cholesterol level, and high triglyceride level) as well age and sex were not compared between these two groups.

### Comparison of clinical parameters between subgroups of stroke patients

To test our hypothesis that “*circadian variation of stroke onset influences early pathogenesis and long-term outcome of stroke among Indians”*, a total of 249 cases with known onset time are categorised by six 4-hour periods. The highest incidence was observed during early morning from 4 am to 8 am [n = 55 (22%); χ^2^ = 5.723, P value < 0.01675) followed by that during the period 8 am to 12 pm; stroke incidence was lowest at night between 12 am to 4 am [n = 20 (8%); χ^2^ = 12.86, P value < 0.00033) [Table 2].

**Table 2:** Comparison of risk factors, initial severity of stroke and cases distribution with respect to stroke onset time.

|  | **0 to < 4** | **4 to < 8** | **8 to <12** | **12 to < 16** | **16 to < 20** | **20 to < 24** |
| --- | --- | --- | --- | --- | --- | --- |
| **Group** | 1 | 2 | 3 | 4 | 5 | 6 |
| Patient number (%) | 20 (8%) | 55 (22%) | 48 (19.3%) | 46 (18.5%) | 43 (17.3%) | 37 (14.9%) |
| Chi-square | **12.86** | **5.723** | 1.431 | 0.73 | 0.117 | 0.467 |
| 95% Confidence Interval | **0.00033** | **0.01675** | 0.23165 | 0.392943 | 0.73254 | 0.4943 |
| NIHSS ± SD (admission) | 12.2±5.67 vs<br>8.73±5.92 | 8.5±4.99 vs<br>8.5±6.22 | 10.53±5.99 vs<br>8.68±6.21 | 7.27±4.87 vs<br>9.49±6.38 | 11.08±8.91 vs<br>8.77±5.43 | 7.19±5.34 vs<br>9.12±6.00 |
| P-Value | **0.01425** | 0.7619 | 0.09894 | 0.1142 | 0.4863 | 0.1714 |
| Recurrent event of stroke | 4 vs 68 | 17 vs 55 | 15 vs 57 | 9 vs 63 | 10 vs 62 | 11 vs 61 |
| P-Value | 0.3638 | 0.7119 | 0.6915 | 0.1254 | 0.3698 | 0.9058 |
| Hypertension (positive) | 18 (90%) vs 176<br>(76.8%) | 46 (83.6%) vs 148<br>(76.3%) | 35 (73%) vs 159<br>(79%) | 32 (69.6%) vs 162<br>(79.8%) | 32 (74.4%) vs 162<br>(78.6%) | 31 (83.8%) vs 163<br>(76.9%) |
| P-Value | 0.0568 | 0.2005 | 0.3919 | 0.1337 | 0.5445 | 0.3538 |
| Diabetes Mellitus (positive) | 13 (65%) vs 90<br>(39.3%) | 19 (34.5%) vs 84<br>(43.3%) | 17 (35.4%) vs 86<br>(42.8%) | 22 (47.8%) vs 81<br>(39.9%) | 17 (39.5%) vs 86<br>(41.7%) | 15 (40.5%) vs 88<br>(41.5%) |
| P-Value | **0.0061** | 0.2658 | 0.3527 | 0.3256 | 0.7888 | 0.9121 |
| Fasting Blood Sugar ± SD<br>(mg/dl) | 186.57±93.40 vs<br>132.01±55.31 | 133.86±62.89 vs<br>134.09±56.84 | 117.97±32.67 vs<br>138.21±62.30 | 122.08±41.14 vs<br>137.0±61.19 | 152.47±79.09 vs<br>128.71±52.23 | 142.67±54.44 vs<br>133.79±59.75 |
| P-Value | **0.01352** | 0.286 | 0.2469 | 0.4832 | 0.1747 | 0.3371 |

MRS: Modified Rankin Scale; NIHSS: National Institutes of Health Stroke Scale; P value <0.05 represented as bold

The comparison of NIHSS score between six groups suggests that immediate severity is highest in patients for subgroup-1 [mean NIHSS score = 12.2; P value 0.01425] *i.e*. cases with night time onset [Table 2]. A statistically significant increased level for fasting blood sugar was also observed in the same subgroup [P = 0.01352] [Table 2]. However, no significant changes for lipid profile and hypertension was identified among the subgroups. No association was also found between circadian variation and occurrence of recurrent events within 6 months from stroke onset.

### Association of time of stroke with cognitive and depression raw scores

Our intergroup analysis on different subdomains of cognition like memory, language, visuospatial skill, executive skill among post-stroke survivors revealed a statistically significant lowering of the raw score representing language problem among night time stroke patients belonging in subgroup-1 [Mean score 28±8.28 vs 33.24±6.53, P value = 0.03027] [Table 3]. Similarly, the patients in subgroup-5 [time of onset: 4 pm to 8 pm] appeared to manifest poor visuospatial skill compared to others [Mean score 4.45±2.91 vs 6.00±3.09, P value = 0.04723] [Table 3]. However, no such difference was identified for memory and executive function between cases when stratified according to time of onset.

**Table 3:** Comparison of cognitive parameters with respect to stroke onset time.

|  | **0 to < 4** | **4 to < 8** | **8 to <12** | **12 to < 16** | **16 to < 20** | **20 to < 24** |
| --- | --- | --- | --- | --- | --- | --- |
| **Group** | 1 | 2 | 3 | 4 | 5 | 6 |
| Patient number (%) | 20 (8%) | 55 (22%) | 48 (19.3%) | 46 (18.5%) | 43 (17.3%) | 37 (14.9%) |
| Memory score | 39.5±9.74vs<br>34.88±11.27 | 36.26±10.25 vs<br>33.78±12.72 | 32.67±14.50 vs<br>34.56±11.85 | 33.81±11.56 vs<br>33.63±13.42 | 32.71±13.85 vs<br>33.94±12.94 | 36.93±8.35 vs<br>33.91±12.69 |
| P-value | 0.1858 | 0.4776 | 0.4722 | 0.8564 | 0.8792 | 0.5349 |
| Language | **28±8.28 vs**<br>**33.24±6.53** | 33.17±6.20 vs<br>31.83±8.73 | 29.87±11.66 vs<br>32.71±7.06 | 33.80±5.94 vs<br>31.59±8.82 | 31.05±8.69 vs<br>32.33±8.21 | 34.77±6.30 vs<br>31.76±8.46 |
| P-value | **0.03027** | 0.729 | 0.5154 | 0.4752 | 0.7176 | 0.3417 |
| Visuospatial skills | 6.29±3.73 vs<br>5.71±3.08 | 6.47±2.85 vs<br>5.56±3.16 | 5.29±3.41 vs<br>5.84±3.02 | 5.85±3.13 vs<br>5.69±3.10 | **4.45±2.91 vs**<br>**6.00±3.09** | 6.57±2.59 vs<br>5.61±3.15 |
| P-value | 0.5889 | 0.2031 | 0.3367 | 0.7231 | **0.04723** | 0.3061 |
| Executive function | 9.86±7.44 vs<br>8.39±5.83 | 9.52±5.91 vs<br>8.22±5.91 | 7.04±5.28 vs<br>8.86±6.00 | 9.23±5.29 vs<br>8.25±6.06 | 6.85±6.29 vs<br>8.84±5.80 | 9.43±6.50 vs<br>8.34± 5.82 |
| P-value | 0.616 | 0.3793 | 0.2036 | 0.3956 | 0.1807 | 0.5767 |
| GDS | **15.38±4.68 vs**<br>**11.65±6.66** | 11.56±7.08 vs<br>12.14±6.56 | 11.93±6.57 vs<br>12.05±6.69 | 10.21±5.35 vs<br>12.49±6.88 | 13.78±7.87 vs<br>11.67±6.35 | 11.56±6.76 vs<br>12.1±6.65 |
| P-value | **0.02007** | 0.5049 | 0.9387 | 0.1454 | 0.2594 | 0.8147 |

P value <0.05 represented as bold

On the other hand, intergroup comparison of GDS values revealed night time stroke as a risk factor for developing depression as long-term outcome [Mean score 15.38±4.38 vs 11.65±6.66, P value = 0.02007] [Table 3].

### Comparison of *CLOCK, BMAL1, PER1* and *CRY1* expression between day and night time stroke patients

Owing to the low number of acute phase patients (n =24) we could able to collect for RNA expression study and influence of diurnal variation in post-stroke cognition and depression [Table 5], here we stratified the diseased subjects into two broad categories namely day time (5 am to 2 pm) and night time onset (9 pm to 2 am) cases. At first, the relative expression of three core clock genes *i.e. CLOCK, BMAL1* and *CRY1* were compared between acute stroke (n = 24) cases and controls (n = 11). A significant difference in all three genes was observed between patients and controls (P < 0.05; Figure 1A as representative).

**Figure 1:**
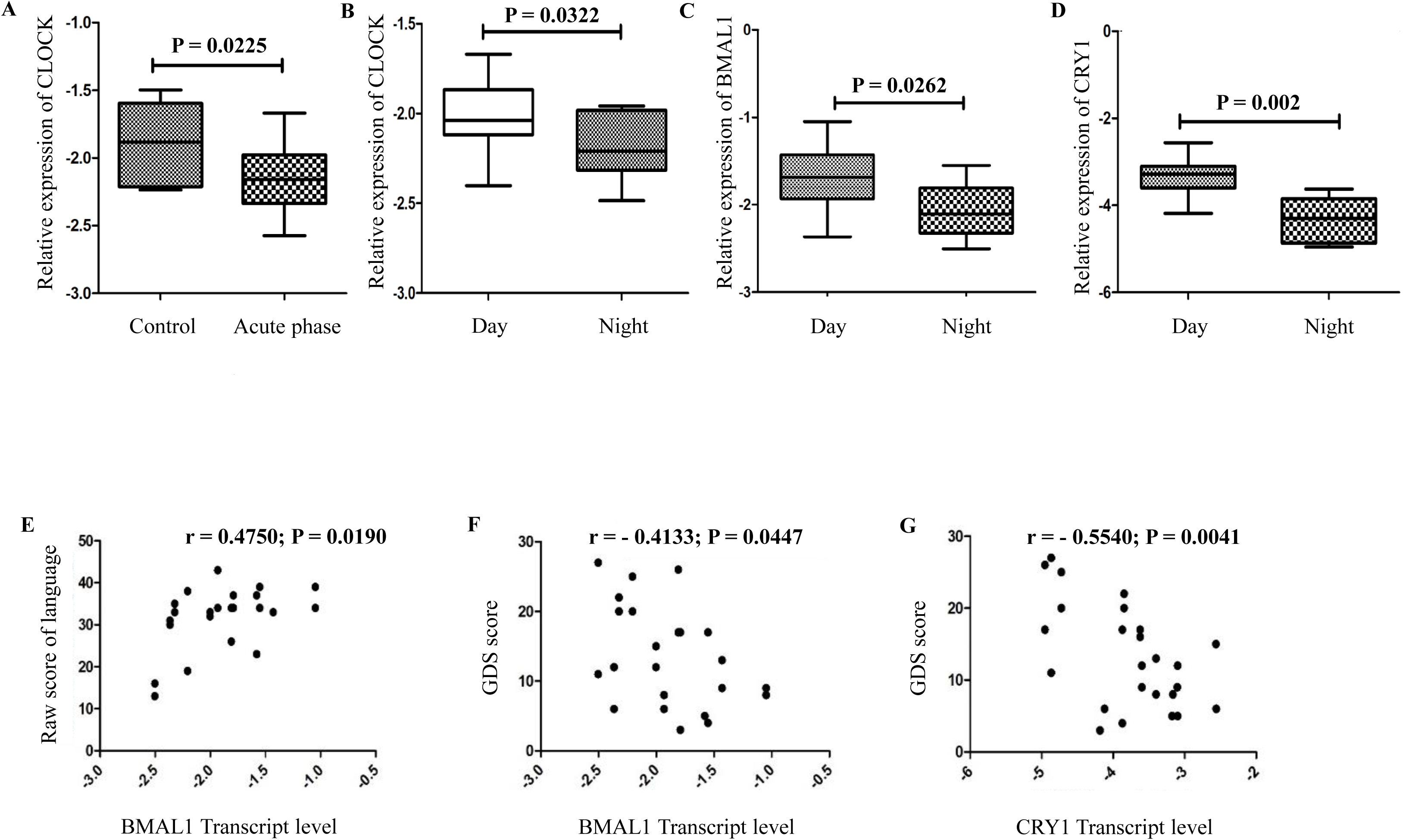
The alteration of circadian gene expression in acute ischemic stroke patients and its correlation with language and GDS score after 6 months. **A:** Quantification of clock genes in PBMC of acute stroke patients (n =24) and controls (n = 11), normalized to 18S using Mann-Whitney U test. **B, C & D:** Quantitative comparison of CLOCK, BMAL1 & CRY1 in PBMC stroke patients in acute phase with different time of onset. Day corresponds to 5 am to 2 pm; night time represents 9 pm to 2 am. Data presented as mean ± standard deviation in all graphs. **E:** A significant correlation of BMAL1 and language score. **F & G:** Significant negative correlation of BMAL1 and CRY1 with GDS score.

Next, to investigate variations in these genes’ expression between day and night cases, the inter group analyses were performed. Here, we found that mean expression of *CLOCK, BMAL1* and *CRY1* genes’ expression is significantly lower for night time cases (Mean log_2_ expression for *CLOCK, BMAL1* and *CRY1* = –2.207 ± 0.0481, –2.066 ± 0.09622, –4.315 ± 0.1638 respectively) than day time patients. (Mean log_2_ expression for *CLOCK, BMAL1, CRY1* = –2.022 ± 0.067, –1.691 ± 0.1242, –3.335 ± 0.1499 respectively) with P values 0.0322, 0.0262 and 0.0002 respectively (Figure 1B–1D).

### Correlation of *BMAL1, CLOCK* and *CRY1* with GDS and language scores

Since the control individuals are non-depressed ones and free of language impairment, here only the stroke patients (n = 24; 12 for day and 12 for night), recruited for RNA analyses were considered for correlation study. For this, the cases were further subjected to follow up study. The mean score for language, GDS scores were 34 ± 4.77, 8.25 ± 3.596 and 29.33 ± 8.37, 15.153 ± 5.817 respectively for day and night cases.

After Spearman r correlation analysis, we demonstrated statistically significant positive correlation between *BMAL1* level and language (r = 0.4750, P= 0.0190). However, for the other two genes’ transcript level no such correlation was observed (Figure 1E).

On the other hand, negative correlation was revealed between GDS score and *BMAl1* (r = – 0.4133, P= 0.0447) and *CRY1* (r = –0.5540, P= 0.0041) (Figure 1F & 1G).

### Distribution of *CLOCK* (rs1801260, rs4580704), *CRY2* (rs2292912) SNVs in different sub groups

On basis of our previous publication and current findings as stated above, here we further evaluated the association of SNVs in core clock genes with circadian variation of stroke onset. Table 4 shows the distribution of rs1801260, rs4580704, and rs2292912 genotypes between cases with different time of onset over the six 4-hour intervals during a day.

**Table 4:** Genotype and Allele frequency distribution of studied polymorphic variants with respect to stroke onset time.

| **rs1801260 (n = 221)** | **Genotype Frequency** |  |  | **Allele Frequency** |  | **P-value** | **Odds ratio (95%CI)** |
| --- | --- | --- | --- | --- | --- | --- | --- |
|  | **TT** | **TC** | **CC** | **T** | **C** |  |  |
| 00:00 – 03:59 (17) | 6 (35.3%) | 9 (52.9%) | 2 (11.8%) | 21 (61.8%) | 13 (38.2%) | 0.7089 | 1.147 (0.558 – 2.36) |
| 04:00 – 07:59 (55) | 20 (36.4%) | 32 (58.2%) | 3 (5.4%) | 72 (65.5%) | 38 (34.5%) | 0.8496 | 0.957 (0.609 – 1.505) |
| 08:00 – 11:59 (43) | 17 (39.5%) | 24 (55.8%) | 2 (4.7%) | 58 (67.4%) | 28 (32.6%) | 0.5543 | 0.859 (0.521 – 1.418) |
| 12:00 – 15:59 (42) | 17 (40.5%) | 18 (42.9%) | 7 (16.6%) | 52 (61.9%) | 32 (38.1%) | 0.551 | 1.161 (0.710 – 1.898) |
| 16:00 – 19:59 (36) | 12 (33.3%) | 17 (47.2%) | 7 (19.5%) | 41 (56.9%) | 31 (43.1%) | 0.1335 | 1.482 (0.886 – 2.477) |
| 20:00 – 23:59 (28) | 15 (53.6%) | 12 (42.9%) | 1 (3.5%) | 42 (75%) | 14 (25%) | 0.0875 | 0.573 (0.302 – 1.085) |
| **rs4580704 (n = 198)** | **GG** | **GC** | **CC** | **G** | **C** | **P-value** | **Odds ratio (95%CI)** |
| 00:00 – 03:59 (16) | 9 (56.3%) | 4 (25%) | 3 (18.7%) | 22 (68.7%) | 10 (31.3%) | 0.460 | 1.344 (0.614 – 2.943) |
| 04:00 – 07:59 (47) | 28 (59.6%) | 17 (36.2%) | 2 (4.2%) | 73 (77.7%) | 21 (22.3%) | 0.386 | 0.7849 (0.454 – 1.358) |
| 08:00 – 11:59 (38) | 18 (47.4%) | 17 (44.7%) | 3 (7.9%) | 53 (69.7%) | 23 (30.3%) | 0.3187 | 1.3239 (0.763 – 2.297) |
| 12:00 – 15:59 (37) | 22 (59.5%) | 11 (29.7%) | 4 (10.8%) | 55 (74.3%) | 19 (25.7%) | 0.9857 | 0.9947 (0.558 – 1.774) |
| 16:00 – 19:59 (33) | 17 (51.5%) | 13 (39.4%) | 3 (9.1%) | 47 (71.2%) | 19 (28.8%) | 0.5378 | 1.2030 (0.668 – 2.166) |
| 20:00 – 23:59 (27) | 19 (70.4%) | 6 (22.2%) | 2 (7.4%) | 44 (81.5%) | 10 (18.5%) | 0.1939 | 0.6176 (0.298 – 1.278) |
| **rs2292912 (n = 208)** | **CC** | **GC** | **GG** | **C** | **G** | **P-value** | **Odds ratio (95%CI)** |
| 00:00 – 03:59 (14) | 7 (50%) | 7 (50%) | 0 (0%) | 21 (75%) | 7 (25%) | 0.1356 | 1.9532 (0.8107 - 4.7057) |
| 04:00 – 07:59 (48) | 15 (31.3%) | 28 (58.3%) | 5 (10.4%) | 58 (60.4%) | 38 (39.6%) | 0.7967 | 1.063 (0.666 – 1.696) |
| 08:00 – 11:59 (38) | 15 (39.5%) | 21 (55.3%) | 2 (5.2%) | 51 (67.1%) | 25 (32.9%) | 0.2709 | 0.7444 (0.440 – 1.259) |
| 12:00 – 15:59 (42) | 15 (35.7%) | 23 (54.8%) | 4 (9.5%) | 53 (60.4%) | 31 (36.9%) | 0.7427 | 0.9204 (0.561 – 1.510) |
| 16:00 – 19:59 (36) | 11 (30.6%) | 21 (58.3%) | 4 (11.1%) | 43 (59.7%) | 29 (40.3%) | 0.728 | 1.0966 (0.653 – 1.842) |
| 20:00 – 23:59 (30) | 8 (26.6%) | 14 (46.7%) | 8 (26.7%) | 30 (50%) | 30 (50%) | **0.0488** | **1.738 (1.003 – 3.014)** |

P value <0.05 represented as bold

**Table 5:** Comparison of cognitive parameters with respect to stroke onset time (night vs day time)

| **Cognitive parameters / stroke onset time** | **16:00 to < 03:59 vs 04:00 to < 16:00** |
| --- | --- |
| Memory score | 34.92±11.75 vs 34.20±12.16 |
| P-value | 0.6588 |
| Language | **30.06±7.63 vs 33.58±5.79** |
| P-value | **0.04745** |
| Visuospatial skills | 5.52±3.08 vs 5.86±3.14 |
| P-value | 0.6153 |
| Executive function | 8.28±6.62 vs 8.60±5.23 |
| P-value | 0.7046 |
| GDS | **13.92±6.69 vs 10.86±6.42** |
| P-value | **0.0215** |

P value <0.05 represented as bold

The patients harbouring ‘CC’ genotype of rs2252912 of *CRY2* showed a mild positive association with late evening time stroke [OR = 1.738; 95% CI = 1.003-3.014; p-value = 0.0488]. However, none of the SNPs studied here could explain the minor and major peaks for occurrence in our study cohort.

## Discussion

The role of circadian clocks’ over physiological processes (sleep cycle, metabolism, immune system, aging learning, and cognition etc.), and circadian pattern of stroke onset, circadian variability of heart rate, blood pressure, blood viscosity, or platelet aggregation is well documented. However, very limited studies stating correlation between PSCI / depression development with time of injury across populations were the major reasons to conduct such study among our study cohort. Here, we provide the direct evidence that time of injury is significantly associated with initial severity and language problem, higher depression in post-stroke survivors among eastern Indians.

In line with an earlier publication in 2023, further we wanted to test whether the sufferings among post-stroke survivors (cognitive impairment and depression) is an outcome of time dependent injury or not which might be associated with circadian gene dysregulation in acute phase [5]. Therefore, in the present study we performed a) transcript level quantification of circadian genes in acute patients with different injury time; b) intergroup comparison (described earlier) for several cognitive subdomain and GDS raw score; and c) correlation between ‘a’ and ‘b’. Our data suggests that, night time stroke makes a person more vulnerable to develop language problem and depression than day time stroke patients. In addition, we also showed a downregulation of *CLOCK, BMAL1* and *CRY1* as underlying mechanism for such observation. Our data is similar to a report identifying *CRY1/CRY2* downregulation to be associated with higher NIHSS score [9] among stroke patients.

Our present data is also concordant to the animal study by Beker et al., in 2018 showing that, night time Ischemic injury (correspond to day time in humans) associated with increased neuronal survival and elevated levels circadian-related proteins including Per1, Bmal1, and Clock proteins resulted in less severe neuronal damage [3]. On the contrary, a recently published study from China in human subjects suggested day time stroke as risk factor for early oxidative stress [10]. Interestingly, the correlation between severity and circadian genes’ expression profile were similar between all these reports.

Furthermore to authenticate our RNA expression data, a correlation analysis was performed between transcript level and clinical raw scores where a positive correlation between *BMAL1* and language and a negative correlation between GDS score and *BMAL1, CRY1* was observed. A number of studies on non-stroke Major Depressive Disorder also found changes in circadian genes’ mRNA level [6]. Therefore, in together, these data vindicate the neuroprotective effect of circadian genes claimed by an earlier *in-vivo* study [11].

Meanwhile, considering the genetic association between variants of clock genes and PSCI among eastern Indians, an attempt was taken to evaluate the same SNVs as risk factors for circadian pattern of onset observed in our cohort. Although an over representation of cases account for day time (Subgroup 2), none of SNVs were found to be differentially distributed in specific subgroups. A missense variant of *Per2* namely, rs35333999, which appeared as a non-informative marker in our population, was found to be associated with later chorotype among Europeans [12] and myocardial infarction among Croatians [13].

Despite of our findings with clinical significance, our study has few limitations including small sample size and correlating biochemical parameters with time of onset. Till date, none of the 3 published work from India correlating TNFα, IL6, IFNγ and C-reactive protein with severity of stroke, have evaluated those as circadian markers [14–16]. Since the prevalence and post stroke outcome vary across different parts of India [17], in our future research we would like to identify most suitable easily diagnosable plasma content (miRNA, exosomal content or proteins) as circadian marker with high specificity and sensitivity.

## Conclusion

Therefore, in conclusion, our study first demonstrates that time of stroke onset has a potential to cause poor cognitive status among eastern Indian stroke patients (Figure 2). Furthermore, circadian genes’ hold a promise to be a potent therapeutic target in order to control depression burden among post-stoke survivors.

**Figure 2:**
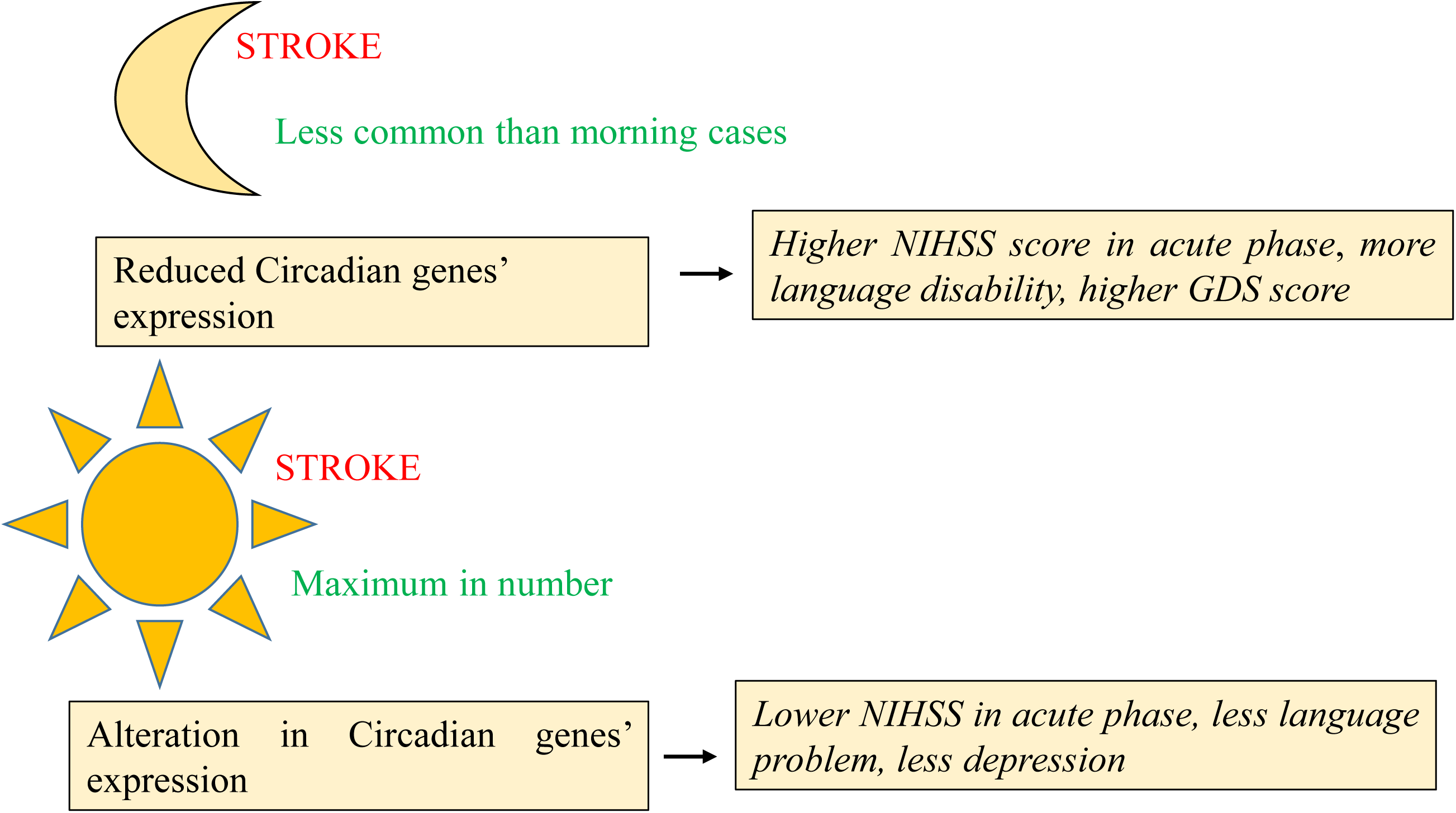
Schematic representation describing night time injury as risk factor for language impairment and higher depression in Post-stroke survivors. The significance level was set at P≤0.05

## Conflict of interest

The authors declare that they have no conflicts of interest.

## Acknowledgements

The authors are thankful to the patients, family members, and healthy individuals who participated in the study.

## Funding

The study has been supported by Post-Doctoral fellowship grants from the Department of Science & Technology, Govt. of India, under Cognitive Science Research Initiative Program to first author DS [DST/CSRI-PDF/2021/12].

## Author Contribution

DS & AB was responsible for the concept, study design, experimental work, and manuscript preparation. AR did the experimental work and help in sample collection along with history. TKB, PK, PSM, JM and KG evaluated patients and blood collection as being clinical collaborators. While, SPH provided the instrumentation facility in S. N. Pradhan Centre for Neurosciences, University of Calcutta and provide his input for manuscript preparation. All authors read the draft, provided their inputs, and agreed on the final version of the manuscript.

## Data availability

The data that support the findings of this study are available from the corresponding author upon request.

## Ethical approval

All procedures performed in studies involving human participants were in accordance with the ethical standards of the National Neurosciences Centre Calcutta, Kolkata, India and Nil Ratan Sircar Medical College & Hospital, Kolkata, India. The Ethics Committees (Institutional Ethics Committee, Nil Ratan Sircar Medical College & Hospital and Institutional Ethics Committee, National Neurosciences Centre Calcutta) of above-mentioned Institutes approved the study protocol. Informed consent was taken as per guidelines of the Indian Council of Medical Research, National Ethical guidelines for Biomedical and Health research involving human participants, India.

## Informed consent

Informed consent from all the participants were received prior to clinical data and sample collection.

## Notes

### Competing Interest Statement

The authors have declared no competing interest.

### Funding Statement

Department of Science & Technology, Govt. of India (DST/CSRI-PDF/2021/12)

